# The Nutritional Status of Primary School Children in Qua’an-Pan Local Government Area, Plateau State

**DOI:** 10.1101/2025.04.09.25325552

**Authors:** Waalmoep Gerald Damar, Morgak James Gonap, Gudzan Sow, Wandayi Emmanuel Amlabu

**Author notes:** Residential address; Opposite ITF office Bukuru Expressway, Jos South Local Government Area, Plateau State, Nigeria.

## Abstract

Nutrition is the fuel that powers an individual and a society at large. Nutrition, if it is given serious attention can also serve as an indicator of the socio-economic wellbeing of a nation. The nutritional status of children is very important as it can lead to deficits in adult life that affect a person’s wellbeing and productivity. This study was carried out to determine the nutritional status of primary school children in Qua’an-Pan LGA, Plateau State, Nigeria. Weight and height of the children were obtained using a bathroom scale and a locally made stadiometer. The age and date of birth of each child was also noted. Hence, the height, weight, date of birth, and date of data collection of each participant was entered into the WHO AnthroPlus software 1.0.4 to obtain Z-scores for the height for age (HAZ) and body mass index (BMI) for age (BAZ). The prevalence of stunting, wasting and overweight/obese was 28.90%, 9.55% and 0.99% respectively. The male children had a significantly higher prevalence of malnutrition than the female children. The high prevalence of malnutrition in Qua’an-Pan Local Government is a worry and it calls for a proper look into the causes of malnutrition among the children and the need to come up with policies and interventions that can help reduce the prevalence of malnutrition among children in the study area.

## Introduction

The processes that involve the intake and the use of food for growth, repair and maintenance of the body is known as nutrition (LeMone *et al*., 2015). Nutrition is understood to be a major component of health and also reflects the economic and social wellbeing of an individual or a population. The overall nutrition and health of a population is affected by changes in body compositions (Galgamuwa *et al*., 2018). Malnutrition is defined as excesses, deficiencies or imbalances in a person’s intake of energy and/or nutrients. Malnutrition takes the form of undernutrition (wasting, stunting and underweight), inadequate vitamins or minerals, overweight, obesity and the resulting diet related non-communicable diseases (WHO, 2018). Undernutrition has been described as one of the most common and persistent health problems in developing countries. More than half of child deaths have been associated with malnutrition (Galgamuwa *et al*., 2018).

Children are at high risk of malnutrition because they need adequate nutrition for growth and development. The effects of malnutrition on the life of children are not only restricted to physical health; it also affects their social and mental capabilities (Gillespie *et al*., 1993; WHO 2016). Sub-Sahara Africa has the highest prevalence of undernutrition with records showing increases in the number undernourished people between 1990-1992 and 2014-2016 (UN, FAO, IFAD and WFP, 2015). Although the United Nations Children’s Fund (UNICEF) has estimated that about 33% of children in Nigeria are stunted, there is no national representative data on the nutritional status of children between the ages of 6-14 years. Institutions such as the Nigeria Demographic and Health Survey (NDHS), the Multiple Indicator Cluster Survey (MICS) and the Nutrition and Health Survey only have national representative data on the nutritional status of children below 5 years and those above 15 years (NDHS 2013, NBS 2014, NBS 2017).

Quaan-Pan Local Government Area is one of the 17 local governments located in Plateau State, Nigeria. It is located in the sourthern part of the state and it is comprised of two major groups of people; the Kofyar and the Goemai people. They are also the major tribes that are found in the Local Government Area. Other tribes resident in the area include the Ibo, Ngas, Tiv, Tarok, Yoruba, Hausa, Mwaghavul and Berom to mention a few.

Anthropometry is a non-invasive and an inexpensive measurement of the human body which can give information about the growth rate of the body. It can tell the nutritional status of the body. Heights, weight as well as the circumference of the head and arms are the main anthropometric measurements carried out on children (Galgamuwa *et al*., 2018; Khair & Morton, 2000).

## Materials and Method

### Study area

Qua’an-Pan (8°48′N 9°09′E / 8.800°N 9.150°E) is made up of eight districts namely; Bwall, Doemak, Dokan Kasuwa, Kwa, Kwalla, Kwande, Kwang and Namu. It is bordered to the north by Bokkos, Mangu and Pankshin LGAs, to the east by Shendam LGA and to the west by Nassarawa State and it covers an area of 2,478 km^2^ with a population of 196,929 people. The people are mostly farmers with some employed as civil servants. They also take part in trading activities (Qua’an-Pan Project Frontiers, 2017). Qua’an-Pan LGA has a tropical climate. It has an average annual temperature of 27.4 °C. Precipitation here averages 1235 mm. The warmest month of the year is April, with an average temperature of 30.3 °C. August is the coldest month, with temperatures averaging 25.8 °C. The driest month is January, with 0 mm of rainfall. Most of the precipitation here falls in September, averaging 249 mm. Throughout the year, temperatures vary by 4.5 °C (Climate-Data.org., 2019).

### Ethical considerations

Ethical clearance was obtained from the plateau state ministry of health and the Local Government Council granted permission for the study to be carried out. The Head teacher of each school that participated in the study gave consent. The parents gave written consent while the pupils that were selected gave oral consent and were informed of their right to opt out of the study at any time without any consequence.

### Study population and design

Sample size for the determination of geohelminth infection was calculated according to the Cochran’s formula (Sarmah, *et al*., 2013)

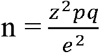

n = sample size

z (standard normal distribution at 95% confidence interval) = 1.96

p (prevalence) = 16.2% (Abah *et al*., 2017)

p = 0.162

q = 1-p = 1 - 0.162

q = 0.838

e = the allowable error, taken as 5% = 0.05

The sample size arrived at was 209 primary school children, however, a total of 806 primary school children were recruited for this study.

Five out of the eight districts found in Qua’an-Pan LGA were randomly selected (by lottery) for the study. A list of all public and private schools in each selected district was obtained from the Department of Education, Qua’an-Pan LGA. The Districts were Kwang, Kwande, Kwalla, Dokan Kasuwa and Bwall. A total of 10 primary schools were selected and two schools were selected from each district. The two selected schools were comprised of a private and a public school and samples were collected from at least 36 children per school.

The cross-sectional study was conducted from the month of September, 2021 to December, 2021. Anthropometric measurements such as the height and weight of the children were determined with a locally made stadiometer and a mechanical bathroom scale (Camry) respectively. The weight of each child was taken with minimal clothing on while the height was taken with each child standing and barefoot. The mean of three weight and height readings for each participant was calculated to minimize error. Readings for the weight and height were taken to the nearest 0.1 kg and 0.1 cm respectively. The age and date of birth of each child was also noted. Hence, the height, weight, date of birth, and date of data collection of each participant was entered into the WHO AnthroPlus software 1.0.4 to obtain Z-scores for the height for age (HAZ) and body mass index (BMI) for age (BAZ) (Masthi *et al*., 2017). Stunting was assessed with HAZ while thinness was assessed with BAZ. Children with HAZ and BAZ below Standard deviation (SD) −2 were classified as stunted, underweight and thin/wasted, respectively. Children with BAZ above SD +2 were described as overweight and obese respectively. Children were classified as malnourished if they had at least one of these two conditions (Galgamuwa *et al*., 2018; Moncayo *et al*., 2017).

Children with apparent disability, sick children and children known to have been sick in the period leading up to the sampling day were excluded from the study. Primary school children or adolescents above the age of 19 years were also exempted from the study.

### Data analyses

Sex, height, weight and the date of birth of each participant was entered into the WHO AnthroPlus software 1.0.4. These measurements were used to calculate the height for age z-score (HAZ) and body mass index for age z-score (BAZ) based on WHO (2007) growth references. The SPSS version 20.0 was used to analyze data. Data was displayed in means, frequencies and percentages. Chi-squared test was used to determine significant differences between variables. P ≤ 0.05 was considered statistically significant.

## Results

### Prevalence of malnutrition among primary school children

Table 1 presents data on the prevalence of malnutrition among primary school children in Qua’an-Pan LGA. Stunting had the highest overall prevalence (28.90%) and it was followed by wasting with an overall prevalence of 9.55%. Overweight had the lowest overall prevalence (0.99%). Chi-square test revealed significant differences in the prevalence of malnutrition indicators (p < 0.01, χ^2^ = 20.99).

**Table 1:** Prevalence of malnutrition among primary school children in Qua’an-Pan Local Government Area.

| District | Number examined | Normal | Stunted (%) | Wasted (%) | Overweight (%) |
| --- | --- | --- | --- | --- | --- |
| Kwalla | 159 | 110 | 39 (24.53) | 9 (5.67) | 1 (0.63) |
| Bwall | 169 | 97 | 57 (33.73) | 15 (8.88) | 0 (0) |
| Kwang | 136 | 83 | 40 (29.41) | 8 (5.89) | 5 (3.68) |
| D.K. | 160 | 95 | 46 (28.75) | 17 (10.63) | 2 (1.25) |
| Kwande | 182 | 103 | 51 (28.02) | 28 (15.39) | 0 (0) |
| **Total** | 806 | 488 | 233 (28.90) | 77 (9.55) | 8 (0.99) |

Keys; D.K.= Dokan Kasuwa, %= percent. $p < 0.01$ , $df = 4$ , $\chi^2 = 20.99$ .

### Prevalence of malnutrition based on districts

Table 1 also shows the prevalence of malnutrition based on the five districts that were sampled in Qua’an-Pan LGA. Kwande district had the highest overall prevalence (43.41%) of malnutrition while kwalla district had the lowest overall prevalence (30.82%) of malnutrition. There was no statistical difference in the distribution of malnutrition among the five sampled districts in Qua’an-Pan LGA (p= 0.80, df= 2, χ^2^= 1.66).

### Prevalence of malnutrition based on sex in Qua’an-Pan LGA

Table 2 presents data on the prevalence of malnutrition among primary school children in Qua’an-Pan LGA based on their sex. The male children had a higher overall prevalence (45.65%) of malnutrition than the female children (32.91%). The difference in prevalence between the boys and the girls was statistically significant when it was analyzed with the chi-square test (p= 0.05, χ^2^= 3.90).

**Table 2:** Prevalence of malnutrition based on sex in Qua’an-Pan Local Government Area.

| Sex | Number examined | Normal | Stunted (%) | Wasted (%) | Overweight (%) | Overall malnourished (%) |
| --- | --- | --- | --- | --- | --- | --- |
| Male | 409 | 220 | 136 (32.85) | 46 (11.11) | 7 (1.69) | 189 (45.65) |
| Female | 397 | 268 | 97 (24.75) | 31 (7.91) | 1 (0.26) | 129 (32.91) |
| Total | 806 | 488 | 233 (28.91) | 77 (9.55) | 8 (0.99) | 318 (39.45) |

Key; %= percent. $p=0.05$ , $df=1$ , $\chi^2=3.90$ .

### Prevalence of malnutrition based on age group

The prevalence of malnutrition among children in Qua’an-Pan LGA based on the three age groups (5-9 years, 10-14 years and 15-19 years) is displayed in Table 3. The 10-14 years age group had the highest overall prevalence (48.54%), it was closely followed by the 15-19 years age group with overall prevalence of 47.17%. The 5-9 years age group had the lowest overall prevalence of malnutrition (21.98%). Chi-square test revealed statistical difference in the prevalence of malnutrition among the three age groups (p= 0.002, df= 2, χ^2^= 12.40).

**Table 3:**
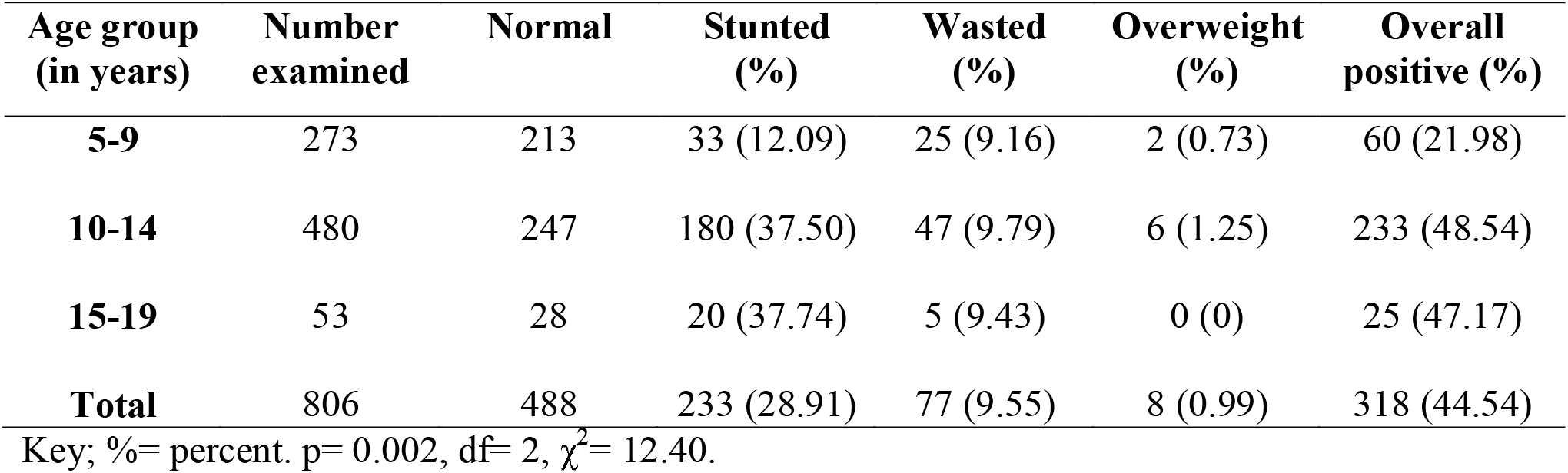
Prevalence of malnutrition based on sex in Qua’an-Pan Local Government Area.

### Prevalence of malnutrition based on ownership of school

The prevalence of malnutrition based on private and public schools is presented in Table 4. The private schools had a higher overall prevalence (40.87%) of malnutrition than the public primary schools (38.13%). The difference in the overall prevalence of malnutrition between the private and public primary schools was not statistically significant (p=0.33, df= 1, χ^2^=0.94).

**Table 4:** Prevalence of malnutrition based on ownership of school in Qua’an-Pan Local Government Area.

| Ownership of school | Number examined | Normal | Stunted (%) | Wasted (%) | Overweight (%) | Overall positive (%) |
| --- | --- | --- | --- | --- | --- | --- |
| Public | 417 | 244 | 114 (27.34) | 42 (10.07) | 3 (0.72) | 159 (38.13) |
| Private | 389 | 203 | 119 (30.59) | 35 (9.0) | 5 (1.29) | 159 (40.87) |
| Total | 806 | 447 | 233 (28.91) | 77 (9.55) | 8 (0.99) | 318 (39.45) |

Key; %= percent. $p=0.33$ , $df=1$ , $\chi^2=0.94$ .

## Discussion

The overall prevalence of malnutrition in this study was 39.45%. It is higher than the prevalence of malnutrition in Jos East LGA (16.48%) and Jos city (35.4%) as reported by Abah *et al*. (2017) and Adedeji *et al*. (2017) respectively. Both study locations are found in Plateau Sate, Nigeria. It is also important to note that both studies were carried out in semi-urban and urban settings. Access to medical care, good water supply and improved socio-economic status of the families in and around the urban areas are factors that help to reduce the prevalence of malnutrition.

Stunting was the most prevalent form of malnutrition recorded in this study. The higher prevalence of stunting in this study agrees with the findings of Abah *et al*. (2017) and Adeomi *et al*. (2021). This is in line with global trends as stunting is known to be the most prevalent form of malnutrition and it is used as an effective indicator of children’s well-being as well as an accurate reflection of social inequality (de onis & Branca, 2016). Overweight had the lowest prevalence as it is recognized as a slow and emerging form malnutrition in the developing world.

There was no statistical difference in the distribution of malnutrition across the five districts that were sampled. This result suggests that malnutrition is evenly spread across the five districts in Qua’an-Pan. The similarity in socio-economic conditions, lifestyle and geographic locations can help to explain the similar prevalence of malnutrition among the five districts that were sampled in Qua’an-Pan Local Government Area, Plateau State.

The male children had a higher and statistically significant prevalence of malnutrition than the female children. This is similar to the findings of Fetuga *et al*. (2010) who reported a higher prevalence among male children than the female children. A reason for the higher prevalence of malnutrition among the male children could be because the male children are allowed to prioritize play and outdoor activities than the female children. The female children are restricted by chores (some African cultures see the female gender as home keepers and are trained to be so from birth) and cooking at home. Excessive play can create an imbalance in the energy obtained through food and the energy expended through play that leads to a deficit. Interestingly, the male children had a higher prevalence of overweight individuals than the female children. This is similar to the work of Igbokwe *et al*. (2018). The small number of overweight (9) individuals reported in this study does not allow a concrete assessment of the condition and it is recommended that more studies be carried out on this condition with a larger sample size to make inferences.

The distribution of malnutrition across the three age groups was statistically similar although the 10-14 years age group had the highest prevalence of malnutrition. Abah *et al*. (2017) reported a similar finding. This is an age of puberty where rapid growth takes place and as a result, quality food is needed by the children to enable the spurt in growth. A deficit in nutrition at this stage can lead to malnourishment and improper growth.

The private primary schools combined had a slightly higher prevalence of malnutrition than the public primary schools but the difference was statistically insignificant. This means that there was an even spread in the prevalence of malnutrition between the public primary schools and private primary schools. This finding is in contrast with the work of Igbokwe *et al*. (2018) in Enugu State (Nigeria) that reported a higher prevalence of malnutrition in the public primary schools. It is an interesting finding because it is believed that most families with high socio-economic status usually send their children/wards to private schools. It would have been logical to suggest that the school feeding program sponsored by the Nigerian government may be the reason for this change in trend since the children attending public schools are fed in school during lunch hours. The school feeding program was active during the course of this research but there were some flaws observed in the program. One major problem with the program was that the food dished out to the children didn’t have the required quality or quantity to meet the nutritional needs of the children. A case in point was when the children were given biscuits as lunch. While biscuits are sweet and nice for children, they do not provide adequate nourishment for the children. A possible reason for the even distribution of malnutrition in this study is the comparable socio-economic status of the people in the area. Most people in the study area are farmers with others engaging in trade and civil service work.

## Conclusion

Malnutrition is prevalent among school children in Qua’an-Pan Local Government Area. Stunting was reported to be the most prevalent form of malnutrition among children in Qua’an-Pan Local Government Area. The sex of the children was observed to be a deciding factor in the prevalence of malnutrition among the sampled children. It is important that the nutritional status of children even before birth is taken seriously as malnutrition may have long lasting effects on their lives and the community at large.

## Data Availability

All data produced in the present study are available upon reasonable request to the authors

## Acknowledgements

Our sincere appreciation goes to the children, parents/guardians, teachers and Head-teachers that participated in this study. Also Mr. Wilberforce Sogotdiel sacrificed a lot to make sure that the researchers had an easy field work. We are most grateful.

